# Excess mortality during the first ten months of COVID-19 epidemic at Jakarta, Indonesia

**DOI:** 10.1101/2020.12.14.20248159

**Authors:** Iqbal RF Elyazar, Henry Surendra, Lenny L Ekawati, Bimandra A. Djaafara, Ahmad Nurhasim, Ahmad Arif, Irma Hidayana, Widyastuti, Dwi Oktavia, Verry Adrian, Ngabila Salama, Imam Hamdi, Adhi Andrianto, Rosa N Lina, Karin D Lestari, Anuraj H Shankar, Raph L Hamers, Guy Thwaites, J. Kevin Baird

**Affiliations:** Eijkman-Oxford Clinical Research Unit, Jakarta, Indonesia; Imperial College London, London, UK; The Conversation Indonesia, Jakarta, Indonesia; LaporCOVID-19, Jakarta, Indonesia; Jakarta Provincial Health Office, Jakarta, Indonesia; Oxford University Clinical Research Unit, Ho Chi Minh City, Vietnam; University of Oxford, Oxford, UK

**Author notes:** **Address for correspondence** Iqbal Elyazar, Eijkman-Oxford Clinical Research Unit. Jalan Diponegoro No 69, Jakarta, Indonesia. 10430.

**Keywords:** COVID-19, SARS-CoV-2, mortality, Jakarta, Indonesia

## Abstract

Excess mortality during the COVID-19 epidemic is an important measure of health impacts. We examined mortality records from January 2015 to October 2020 from government sources at Jakarta, Indonesia: 1) burials in public cemeteries; 2) civil death registration; and 3) health authority death registration. During 2015-2019, an average of 26,342 burials occurred each year from January to October. During the same period of 2020, there were 42,460 burials, an excess of 61%. Burial activities began surging in early January 2020, two months before the first official laboratory confirmation of SARS-CoV-2 infection in Indonesia in March 2020. Analysis of civil death registrations or health authority death registration showed insensitive trends during 2020. Burial records indicated substantially increased mortality associated with the onset of and ongoing COVID-19 epidemic in Jakarta and suggest that SARS-CoV-2 transmission may have been initiated and progressing at least two months prior to official detection.

**Article summary line:** Analysis of civil records of burials in Jakarta, Indonesia showed a 61% increase during 2020 compared to the previous five years, a trend that began two months prior to first official confirmation of SARS-CoV-2 transmission in the city.

## Introduction

The ongoing COVID-19 epidemic exerts negative impacts on the health and wellbeing of Indonesians (*1*). Health authorities confirmed the first two SARS-CoV-2 cases on 2 March 2020. Tracing revealed contact with a Japanese national who had tested positive for the virus in Malaysia after visiting Jakarta in early February (*2*). Even earlier transmission of SARS-CoV-2 is suspected on the basis of direct travel links to the pandemic epicenter of Wuhan, China (*3*) and findings from mathematical modeling (*4*). Once introduced the virus quickly spread to all 34 provinces of Indonesia (*5*).

As of 12 November 2020, Indonesia had detected almost 612,000 SARS-CoV-2 infections among nearly 4.2 million people tested out of a total population of 270 million, the world 4^th^ most largest country (*6*). Among the top 30 confirmed COVID-19 national case tallies, Indonesia has the second lowest per capita testing rate at 15 thousand/million population vs. top-testing United Kingdom at >600 thousand/million population (*7*). This shortfall in testing capacity likely comes with very significant underreporting of infection and its consequences (*8*). Almost 19,000 Indonesians have died following a confirmed diagnosis of SARS-CoV-2 infection. That number is inevitably affected by testing capacities (*9*). Some patients may die without testing, whether in hospital or at home, or testing may be falsely negative (*10*). Indirect mortality linked to COVID-19 may arise from impaired access to healthcare or the impacts of social isolation and economic hardships (*11*). Test-confirmed COVID-19 deaths likely underestimates the number of deaths attributable to the pandemic, especially where testing capacities are limited as in lower and middle income countries (LMIC) like Indonesia (*12*).

Assessing mortality caused by the historic COVID-19 pandemic is a global health imperative (*13*). Estimation of deaths in excess of what had been normal before the pandemic has been used as a measure of its severity (*14,15*). These excess death statistics, which are not influenced by differential testing capacities, may be used to compare across- and within-country COVID-19 impacts (*16*). Excess death intelligence may be leveraged to formulate control strategies, prioritize prevention and treatment resources, target impacts evaluation, and research (*17*). How death is recorded and reported varies widely among and within nations, and present challenges, but valuable inference can be gained (*14,18*).

The present study aimed to assess excess mortality that occurred during the first ten months of SARS-CoV-2 epidemic within the city of Jakarta, Indonesia. We examined records of deaths and burials through several official sources, assessing the comparative strengths and weaknesses of each. That assessment guides on-going efforts to estimate excess mortality in other large urban centres of Indonesia as a means of measuring the health impacts of COVID-19 epidemic occurring in this populous LMIC.

## Methods

### Study design

We employed a retrospective population records-based observational study design. Three distinct datasets were assessed, each originating from official government sources: 1) civil administrative records of burials; 2) civil authority death registrations; and 3) health authority death registration records. In all three datasets, we examined records from 1 January 2015 to 31 October 2020.

### Study location

The capital city of Jakarta administered as the Province of Daerah Kota Istimewa (DKI) is situated on the northwestern coast of the Java Island in tropical Southeast Asia. DKI Jakarta consists of five administrative cities composing 46 sub-districts, 267 neighborhoods, and 2.7 million households (*19*). Daytime temperatures range between 23°C and 35°C during six month wet and dry seasons. Approximately 11 million people live within a 664 km^2^ land area (15,900 people per km^2^). Its population growth rate is 1.19% per year. The population age profile is 25% <15 years old, 14% aged 15-24 years, 49% 25-54 years, 8% 55-64 years and 5% >65 years of age. There are 88% of residents practicing Islamic religion, 13% Christian, 4% Buddhist, and 0.2% Hinduism. The average monthly individual income was approximately USD 310. Nearly 14,100 medical doctors and 27,100 nurses provide medical services at 189 public and private hospitals and 315 primary health centers (*19*). DKI Jakarta is highly mobile and connected, with nearly 12 million registered motor vehicles (predominantly motorcycles); government-operated rail and bus lines service within-city and beyond to surrounding satellite cities with 336.8 million and 264 million passengers per year, respectively; in 2019 Jakarta’s two airports received 7.3 million international and 19.5 million domestic passengers. During 2020 all of these transportation services experienced very substantial declines in these numbers due to voluntary or compulsory limits to access.

### Data sources

#### Civil administration burial registrations

The Jakarta Burials Office compiles funeral data from 75 public cemeteries (*20*) occupying an area of 6 km^2^. Citizens apply to this office for grave allocation and usually receive it within a few hours, an extraordinary efficiency driven by the Islamic custom of burial having to occur before the sun sets on the day of death. These registered burials do not include several other sources of human remains disposal within the city; cremations, burial in private cemeteries, or burial/cremation outside of Jakarta. No records of those activities are kept by the administration. The research team extracted all individual funeral records between 1 January 2015 and 30 October 2020. Each record contained name of the deceased, date of death and funeral, religion, and location of burial of remains. These were entered into a relational database (Microsoft Access) and checked for entry errors and repeat reporting of same death (requests for grave allocation sometime inadvertently placed by both the family and health authorities).

The National Guideline of Prevention and Control for SARS-CoV-2 infections and Guideline of Funeral Services for COVID-19 deaths recommend health and mortuary workers managing the corpse apply the same precautionary measures to either a COVID-confirmed or - probable death (*21-23*). Because management of remains is recorded as conforming to “COVID-19 protocol”, classification as a probable COVID-19 death may occur with either unexamined, inconclusive, or awaited laboratory results (*24*). As the COVID-19 epidemic in Jakarta expanded in March 2020, two public cemeteries were designed to receive the remains of those deceased with a confirmed or suspected COVID-19; Pondok Ranggon and Tegal Alur.

#### Civil authority death registrations

The civil authorities require registration of deaths with the DKI Jakarta Population and Civil Registration Office, a laborious process passing through several lower levels of administration. Relatives of the deceased begin with submitting a death registration form to a neighborhood authority, which is in turn submitted through two higher administrative authorities before registration at the DKI Jakarta office. This process may take weeks to complete and usually involves legal needs required for specific purposes such as insurance, taxation, or estate disposition. The agency has little or no means of checking or enforcing compliance to registration. The research team entered aggregated death statistics extracted from documents published by Jakarta Population and Civil Registration Office between January 2015 and October 2020 to a standardized database. Documents contained all-cause deaths by month, gender, and neighborhood location (*25,26*). The records available to us contained only date of registration of death rather than date of death.

#### Health authority death registrations

The DKI Jakarta Health Office conducts surveillance of deaths occurring within the health facilities it operates. These authorities issue an official medical cause of death certificate using the International Classification of Disease-10 (ICD-10). Families of those who die outside of health facilities may also apply to these authorities for that certificate. Issuing the certificate is typically driven by the legal needs of survivors of the deceased. The process does not appear to be initiated in all instances of death. We extracted digital death records between 1 January 2015 and 30 October 2020 from the health-facility disease surveillance system from the Jakarta Health Office. With onset of known COVID-19 cases in early March 2020, the same office immediately began classifying records of death associated with laboratory-confirmed COVID-19, which we extracted up to 30 October 2020.

## Statistical Analysis

We calculated the normal baseline level using the average of burial and deaths between 2015 and 2019. A 95% confidence interval was calculated for each particular week or month. We calculated excess burials or deaths by calculating the number of burials or deaths above the weekly or monthly baseline level (*27-29*). We assessed the number of excess burials that could be explained by SARS-CoV-2 infection. Statistical analysis was performed using the Stata 12.0 program (StataCorp, College Station, TX, USA).

## Results

Figure 1 illustrates analysis of weekly burials reporting for DKI Jakarta for the five years preceding onset of the COVID-19 epidemic compared to the same events during 2020. During 2015-2019, burials averaged about 600/week, and remained at that level until mid January 2020 with a relatively modest surge to nearly 800 burials that declined to normal range by early March 2020. During March to August 2020 burial activity surged to a relatively stable 1,000 burials per week. In early September, burials surged to over 1400/week but by early November declined to about 1,000/week. The sum of burials activity for the first 10 months of 2020 (January to October) was 42,460 whereas the average sum for the same months in 2015-2019 was 26,342 burials. We thus observed a 61% increase in burials during 2020.

**Figure 1.**
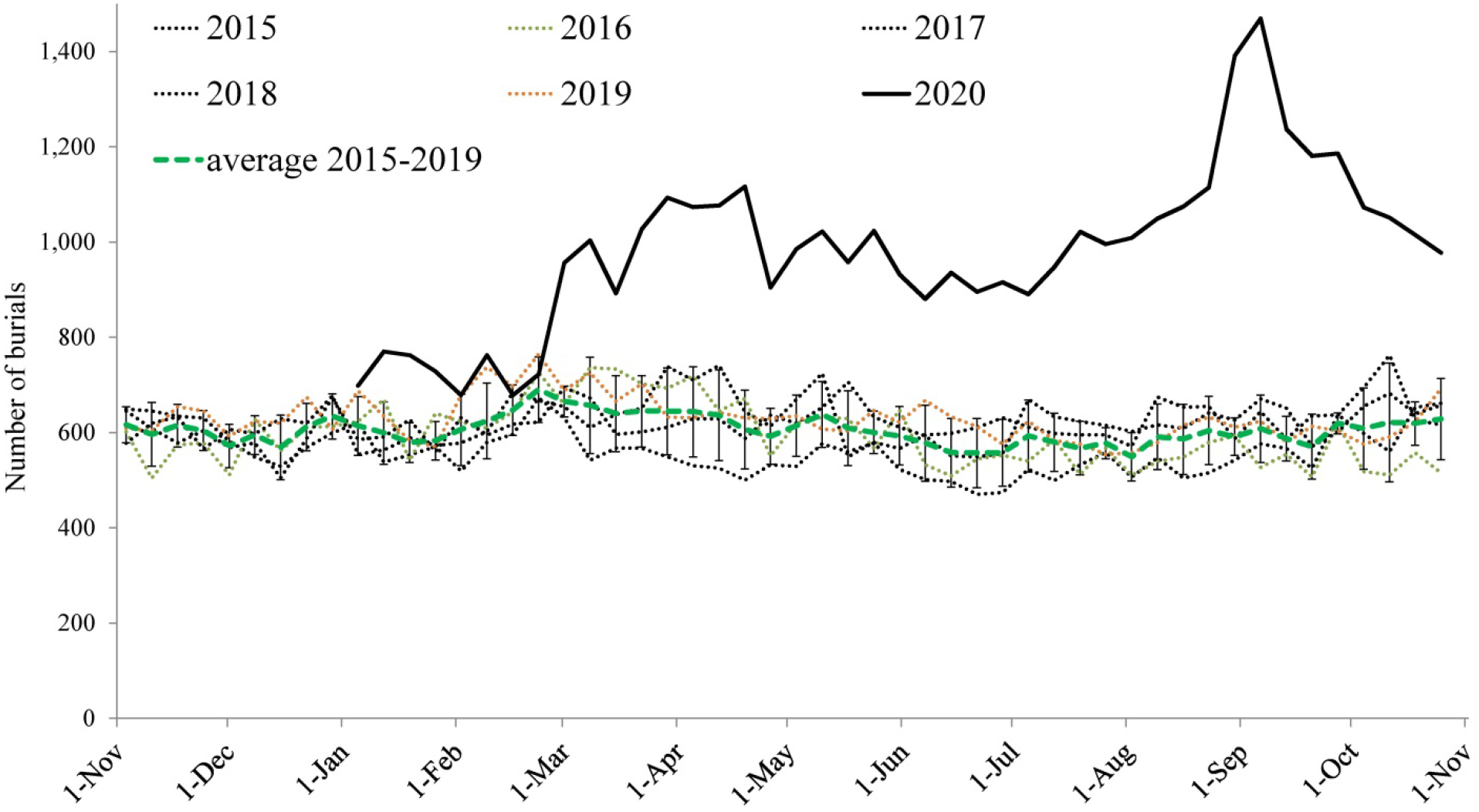
All-cause weekly burials reported between 2015 and 2020. Colored dash lines represent the average number of all-cause weekly burials between 2015 and 2019 (grey solid lines for each year). Solid black line represents the number of all-cause weekly burials in 2020.. A 95% confidence interval is plotted for each all-cause weekly burials during 2015-2019.

Figure 2 illustrates weekly burials as total excess above normal and the subset of burials classified according to COVID-19 protocol. The excess in burials began in early January 2020, rising to nearly 200 by mid-month but declining to nearly normal activity by mid-February. Later than month, burial activity sharply increased and was followed in early March by the very first COVID-19 protocol burials after the first confirmed cluster of infections on 2 March 2020. Thereafter, the trends in excess burials followed the same pattern of COVID-19 protocol burials. During the months of March to November 2020, 16,118 excess burials occurred, with 7,795 (48%) of those by COVID-19 protocol.

**Figure 2.**
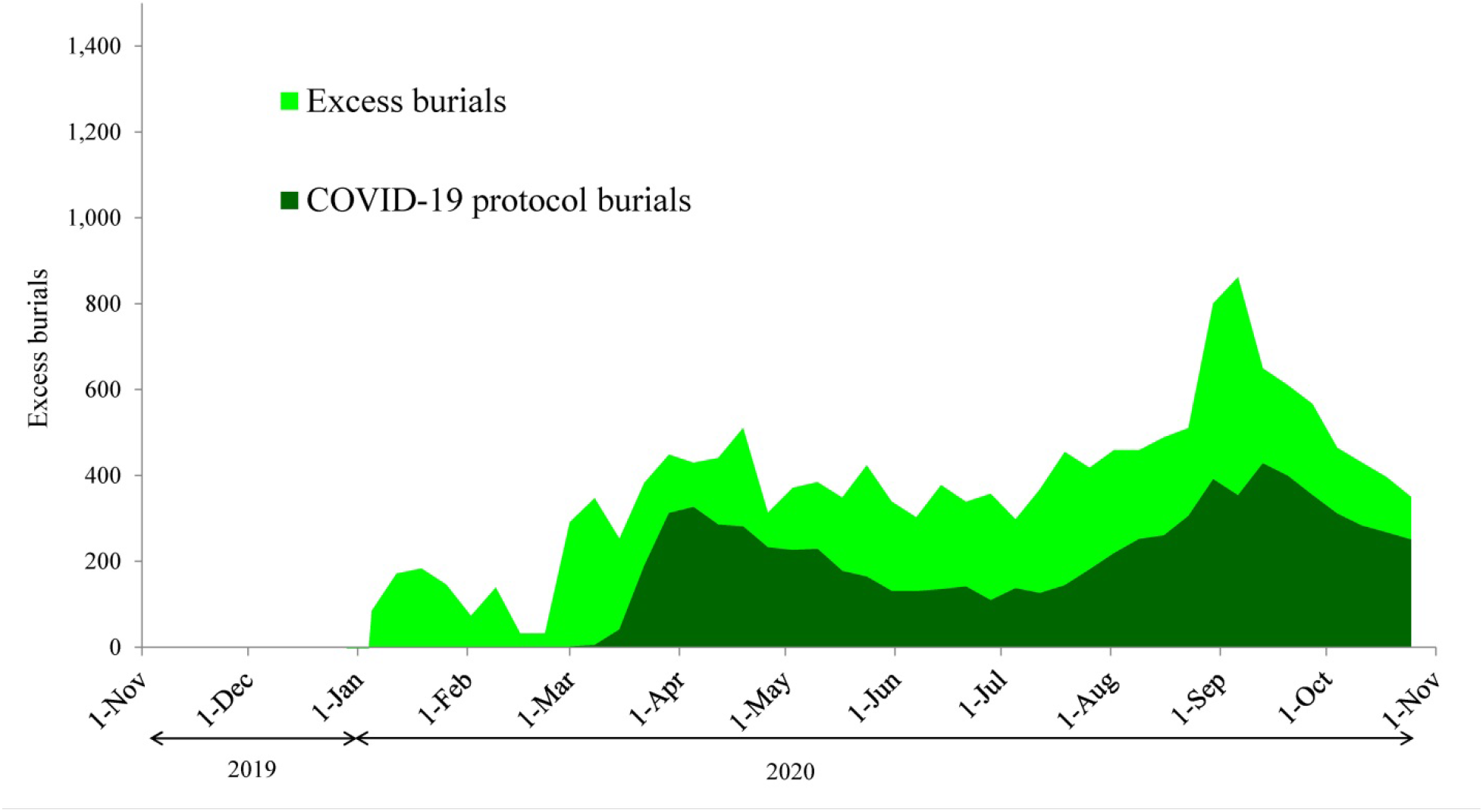
Excess burials estimates during the onset of COVID-19 epidemic in Jakarta between March and October 2020. Light green area represents weekly registered burials. Dark green area represents burials by COVID-19 protocol.

Figure 3A illustrates monthly death registrations with the civil authorities of Jakarta between January 2015 and October 2020. The monthly totals excluding 2020 were stable at around 5,000 registrations. During March, April, and May those registrations declined significantly but then rose sharply in late May and early June to high levels sustained through July and August. Figure 3B illustrates monthly registrations of deaths by the health authorities of Jakarta from January 2015 to October 2020. These registrations during 2020 fell within the norms, with the exception of the very slight elevation in January 2020 and the diminished reporting for October 2020.

**Figure 3.**
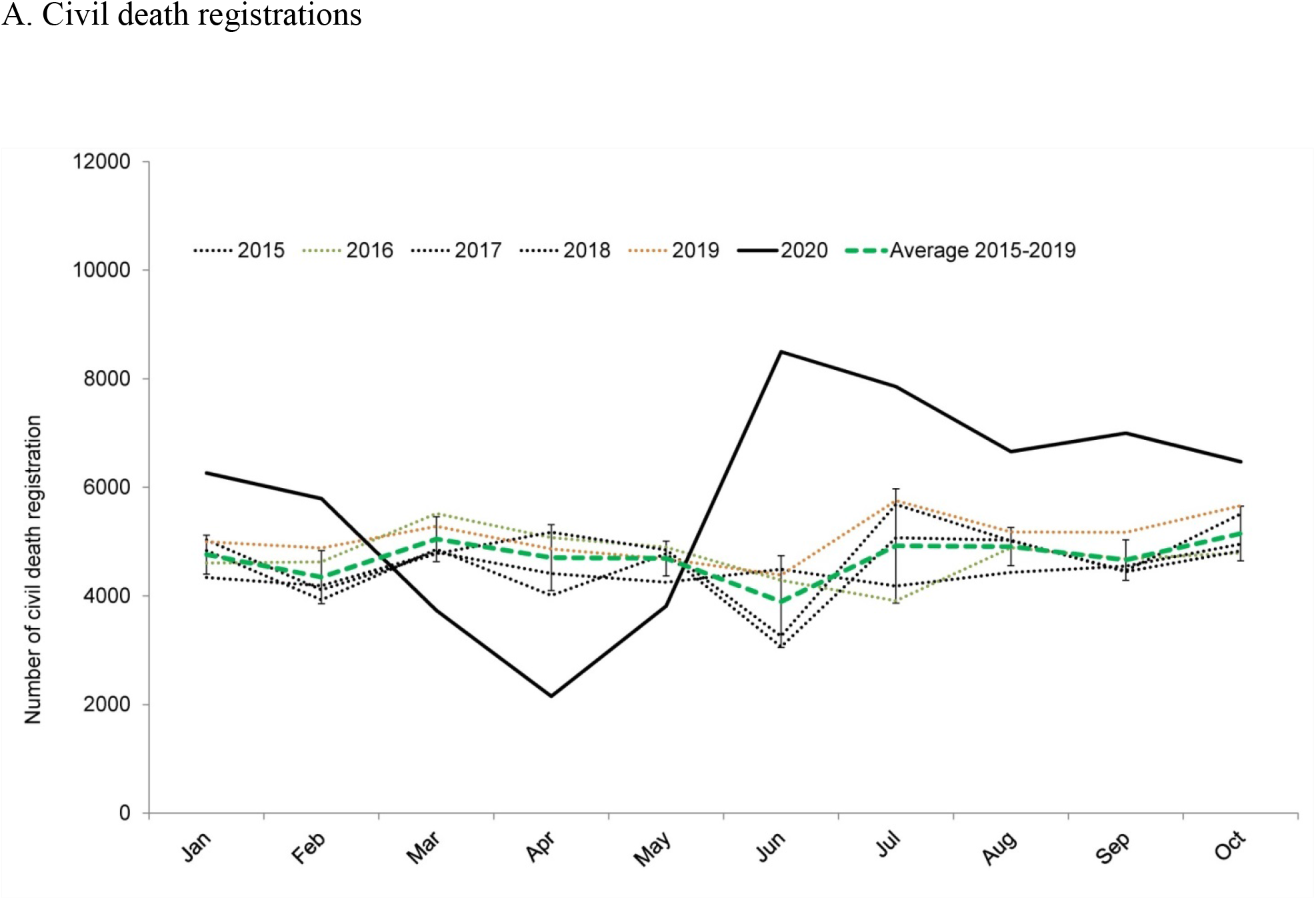
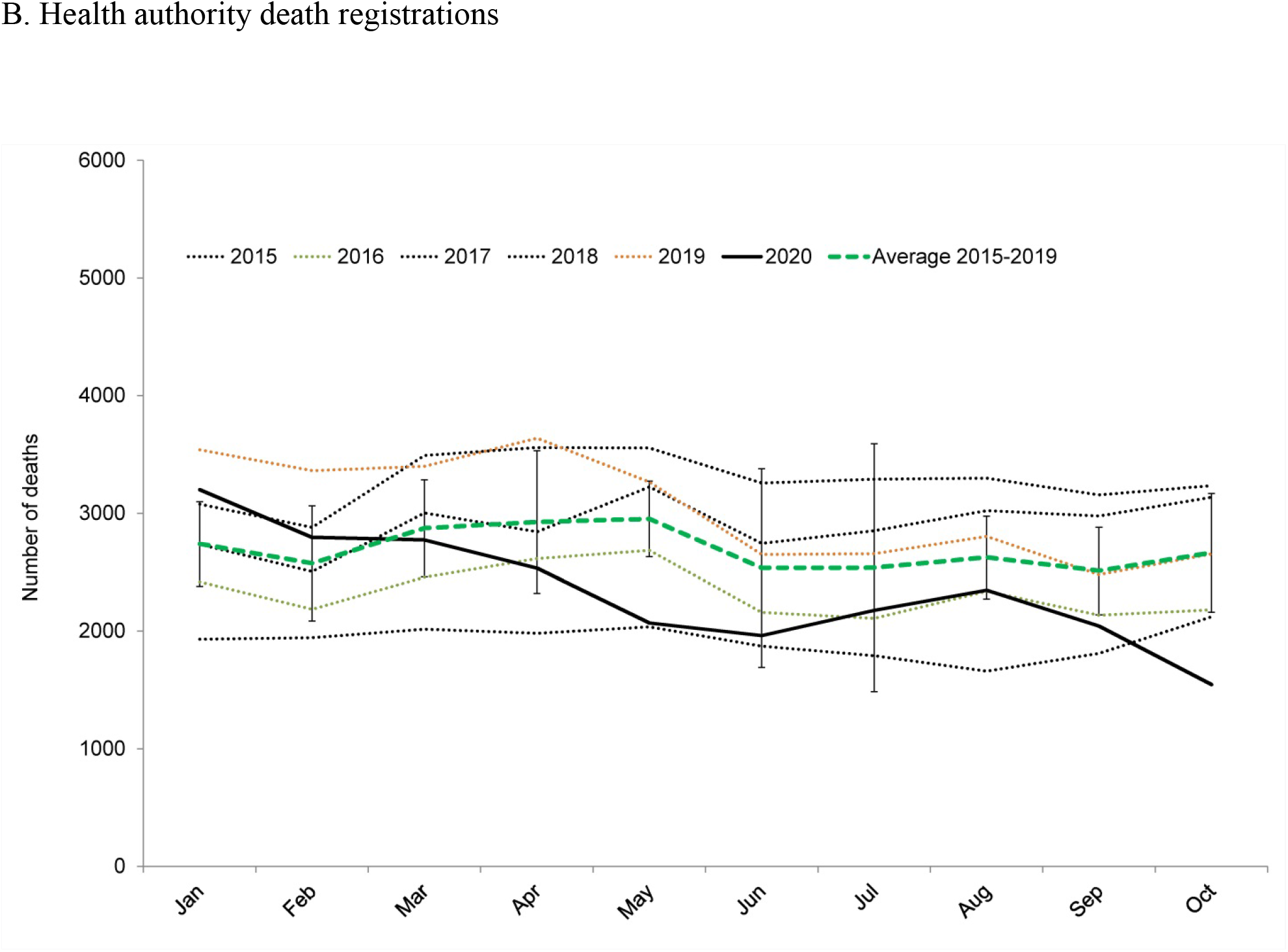
Monthly numbers of deaths registered by civil authority death registrations (A) and by health authority death registrations (B) from January 2015 to October 2020.

## Discussion

Trends in civil records of burials activities in the public cemeteries of Jakarta during 2015-2020, indicate a 61% increase during COVID-19 epidemic. The absolute excess of 16,118 funerals during 2020 does not capture all excess deaths, but only those of residents seeking burial in the public cemeteries operated by the civil authorities. Numbers of funerals and cremations of residents of Jakarta occurring at private facilities or outside of the city are not known. All cause mortality values are thus not known, but the analysis of these burials suggests a 61% increase associated with the ongoing epidemic. News media sources described 55% excess burials activity in Jakarta and from 12% to 270% in other large cities around the globe during 2020 (*28,30,31*). The trend in Jakarta differs from those other cities in at least one important aspect, whereas others experienced peaks in estimated mortality followed by sharp or gradual declines, mortality in Jakarta has not yet peaked.

All burials according to COVID-19 protocol were captured by the city mandate to use public cemeteries designated for that purpose. Those burials accounted for 48% of the excess registered burials representing an unknown proportion of all-cause mortality in the city. We thus cannot know the direct contribution of SARS-CoV-2 infection to that mortality or the indirect casualities due to the broader clinical, social, and economic impacts of the epidemic. Nonetheless, trends in records of burial arrangements provided objective evidence and measure of a sustained surge in mortality associated with COVID-19 transmission.

Burials activity in Jakarta increased sharply two months prior to the first laboratory confirmation of SARS-CoV-2 infection in Indonesia, suggesting probable establishment of transmission in Jakarta during December 2019. Other analyses also pointed to virus introduction to Indonesia (mostly via international arrivals from Wuhan, China to Jakarta or Bali) during late 2019 (*3*). The apparent long duration between introduction and detection points to key weakness of systems of public health surveillance and laboratory capacities in Jakarta, as seen elsewhere (*32,33*). Likewise, trends in registration of deaths through civil or health authorities showed small or delay changes in relation to onset and progression of the epidemic in Jakarta. The data from civil authorities recorded date of registration of death rather than date of death, explaining the otherwise implausible sharp drop in registrations during the first three months of the epidemic. A lockdown period imposed by the city authorities (15 March to 5 June 2020) sharply limited access to registration services. The registration of deaths in health facilities showed almost complete insensitivity to rising mortality in the city, raising concerns regarding its utility as a means of death registration.

Our findings suggest SARS-CoV-2 probably began circulating in Jakarta during December 2019, but remained undetected until early March 2020. The infection provoked a sustained 61% surge of excess burials in the city, about one half of which were suspected or diagnosed as due to COVID-19. Poor capacities of laboratory diagnosis and of death surveillance impose uncertainty in both the characterization and management of this ongoing health crisis in Jakarta. As reported by others, some evidence suggests that non-therapeutic interventions implemented by the civil authorities, and broad acceptance and compliance to those, averted far higher numbers of deaths in the city (*4*). The public burials data described here may provide a useful measure of mortality impacts and of the success of interventions against the epidemic.

## Data Availability

The datasets generated and analyzed during the current study are available from the corresponding author on reasonable request

## Acknowledgments

This study was approved by the Health Research Ethics Committee of the National Institute of Health Research and Development, Ministry of Health Indonesia (LB.02.02/2/KE.554/2020). This work was funded by the Wellcome Trust, UK (106680/Z/14/Z). The funder of the study had no role in the study design, data collection, data analysis, data interpretation, or writing of this paper. The team thanks all data contributors for their contributions in providing data to this study. We thank to Muhammad Akbar and Alisha Nefertity for extracting and compiling burials data records, Agus and Windy for arranging contract and funding management.

## Disclaimers

The opinions expressed by authors contributing to this journal do not necessarily reflect the opinions of the institutions with which the authors are affiliated.

## About the Author

Iqbal Elyazar is a biostatistician who leads the Geospatial Epidemiology Program at the Eijkman-Oxford Clinical Research Unit. He has focused on biostatistics, disease surveillance, and geospatial epidemiology for 20 years. He received a Wellcome Trust Fellowship of Public Health and Tropical Medicine in 2012.

